# A Novel Gene ZNF862 Causes Hereditary Gingival Fibromatosis

**DOI:** 10.1101/2020.04.27.20080804

**Authors:** Juan Wu, Dongna Chen, Hui Huang, Ning Luo, Huishuang Chen, Junjie Zhao, Yanyan Wang, Yang Ren, Teng Zhai, Weibin Sun, Houxuan Li, Wei Li

## Abstract

Hereditary gingival fibromatosis (HGF) is the most common genetic form of gingival fibromatosis which is featured as a localized or generalized overgrowth of gingivae. HGF is genetically heterogeneous and usually be transmitted as an autosomal-dominant inheritance pattern, also can be as autosomal-recessive or occur sporadically. Currently only two genes (*SOS1* and *REST*), as well as four loci (2p22.1, 2p23.3-p22.3, 5q13-q22, and 11p15), have been identified as associated with HGF in a dominant inheritance pattern. Here we report thirteen individuals with autosomal-dominant non-syndromic HGF from a large four-generation Chinese family. Whole-exome sequencing followed by further genetic co-segregation analysis was performed for the family members across three generations. A novel heterozygous missense mutation (NM_001099220.3: c.2812G>A) in zinc finger protein 862 gene (*ZNF862*) was identified, and it is absent among the population as reported from the Genome Aggregation Database, Exome Aggregation Consortium (ExAC) and 1000 Genomes. ZNF862 is a predicted intracellular protein which function is not yet identified, as a zinc finger protein it may be involved in transcriptional regulation. ZNF862 is expressed ubiquitously across tissues, it may play various roles under different physiological condition. Here for the first time we identify the physiological role of *ZNF862* for the association with the HGF trait.

---

Gingival fibromatosis (GF) is a rare disorder which is characterized by a benign, nonhemorrhagic, localized or generalized fibrous enlargement of free and attached gingivae with slow progression. GF could be complicated by epilepsy, hypertrichosis, and mental retardation^1,2^ or it can develop as a part of syndromes like Cowden’s syndrome,^3^ Zimmerman-Laband syndrome,^4^ Cross syndrome,^5^ Rutherford syndrome,^6^ Ramon syndrome,^7^ Jones syndrome,^8^ Costello syndrome,^9^ Ectro-amelia syndrome,^10^ and hyaline fibromatosis syndrome;^11^ it may also be caused by poor oral hygiene related gingival inflammation, puberty, pregnancy or a side effect of the common medications such as calcium channel blockers (nifedipine and verapamil);^12^ usually it betrays as an isolated condition as non-syndromic hereditary gingival fibromatosis (HGF).^13^ HGF is the most common genetic form of GF and usually transmitted as an autosomal-dominant inheritance pattern. Nevertheless, sporadic cases and autosomal-recessive inheritance pedigrees has also been previously described.^14^

As reported previously, to date, four loci (2p22.1 [MIM: 135300], 5q13–q22 [MIM: 605544], 2p23.3–p22.3 [MIM: 609955], and 11p15 [MIM: 611010]) have been identified associated with HGF;^15–18^ besides a heterozygous frameshift mutation in *SOS1* (MIM: 182530) has been identified as causative for the autosomal-dominant HGF in a Brazilian family;^19^ and protein truncating mutations in *REST* (MIM: 600571) have been reported as the genetic cause of an autosomal-dominant inheritance pattern HGF across three independent families.^20^ The estimated incidence of HGF is 1 per 175,000 of the population^21^, and equal between males and females.^22,23^ Gingivectomy can be applied to patients with HGF due to the aesthetic and functional concern, whereas the recurrence of hyperplasia is relatively high potentially owing to the underlying genetic predisposition.^24,25^ Therefore, it is important to explore the genetic causes and etiology accounting for HGF with expectations of the precise genetic diagnosis and of the potential gene therapy development, to help individuals improve their lives. We report a large Chinese family with thirteen affected individuals clinically diagnosed with non-syndromic HGF, and with twelve unaffected family members. Genetic analysis of this large family led to the identification of a heterozygous mutations in a novel gene *ZNF862*.

This four-generation pedigree with HGF was shown in Figure 1A. Visual inspection indicated severe or mild generalized GF in all affected members, whereas not in any unaffected member, from this family. No individuals in this family was ever exposed to the medication that may result in GF according to our investigation. No other congenital abnormality has been found for all of the family members. The clinical data of all individuals in this family are summarized in Table 1, and clinical photos of one control and three patients from each generation are available in Figure 1B. From a clinical perspective, the proband (IV-2 in Figure 1A) had the mild recurrence of hyperplasia after the gingivectomy, and the patient II-2 had an additional clinical finding (hypertension). This study was approved by the Institutional Review Board on Bioethics and Biosafety of BGI (BGI IRB No. 19059) and the informed consent was signed by every participant. Clinical investigation was performed in accordance with the Declaration of Helsinki.

**Figure 1.**
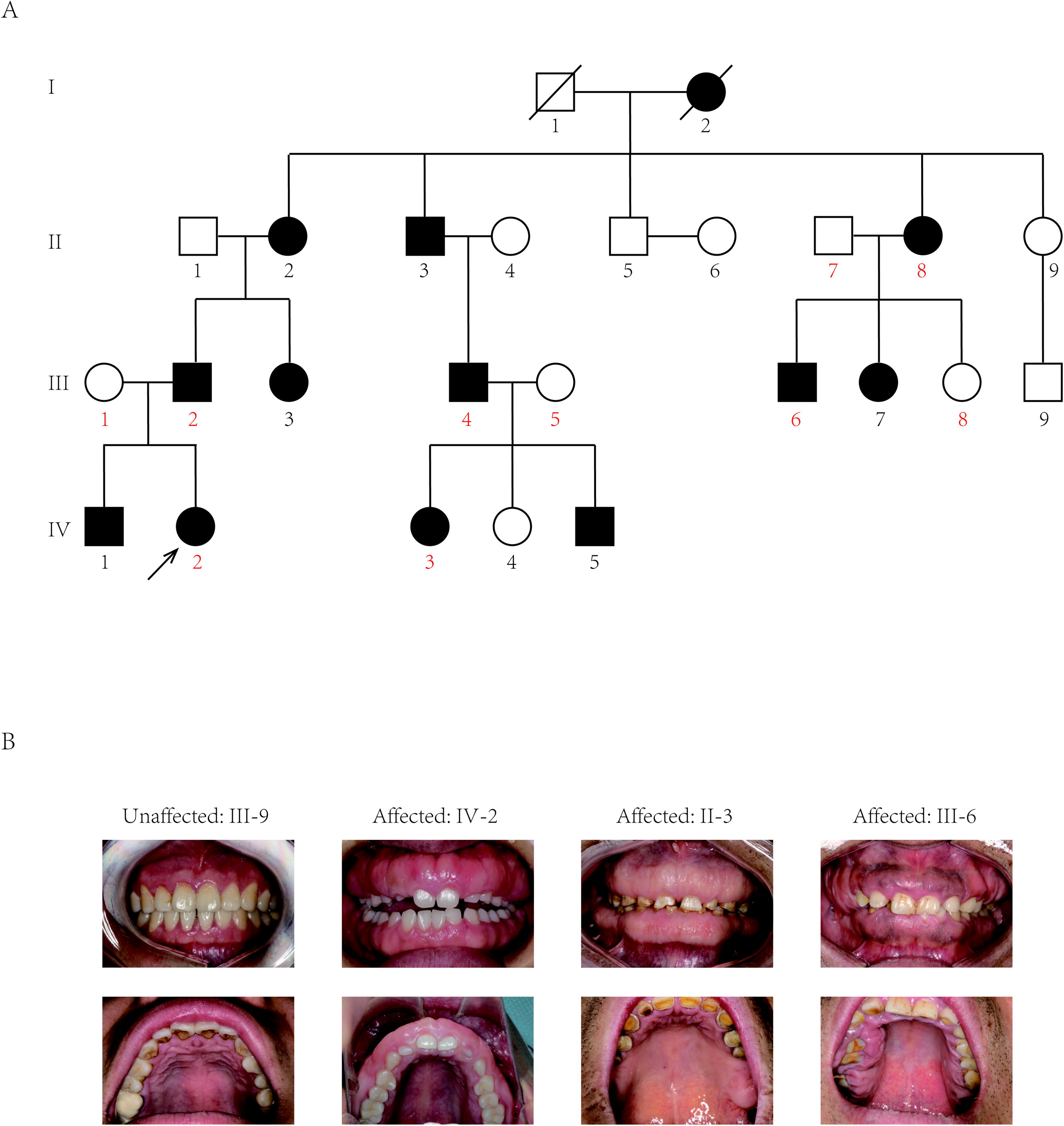
Pedigree and Gingival Photographs of the Individuals. (A) The black arrow indicates the proband. The affected individuals are indicated with black-filled boxes in this family. (B) The gingival overgrowth is revealed by intraoral examination in affected members compared with in the unaffected member in this family.

Whole-exome sequencing (WES) was performed for probands (IV-2 in Figure 1A) and nine other members (numbers indicated in red color in Figure 1A) in the family according to previously described protocols.^26^ The samples achieved 95% of the targeted exome bases covered to a depth-of-coverage of 20× or greater, and totally average sequencing depth for each sample was 100× or greater. Sequencing reads were aligned to the human genome reference sequence (hg19) through the in-house programs. Genome Analysis Toolkit (GATK) was used to call single nucleotide variants (SNVs) and indels; and Annotation of Genetic Variants (ANNOVAR) was used for the annotation. A custom Perl script was used for retrieving the variants corresponding to the inheritance model of this family and that do not appear in unaffected individuals.

After analyzing all SNVs and indels for each gene including the promoter region, exons, splicing sites, introns, and UTR regions, only three variants *ATP7B* (c.3403G>A), *CDADC1* (c.83–13G>T), *ZNF862* (c.2812G>A) were identified to be co-segregated with the phenotype among the ten family members who underwent WES. To further screen and validate the potential disease-causing variants co-segregation with the phenotype in this pedigree, conventional PCR was performed for sanger sequencing evaluation. Eventually, only *ZNF862* (c.2812G>A), no any other variant, was validated to be co-segregated with the phenotype in all twenty-three alive members in this family, as is shown if Figure 2A. This variant has been absent among the large population according to the report of Exome Aggregation Consortium (ExAC) database, 1000 Genomes, and Genome Aggregation Database.

**Figure 2.**
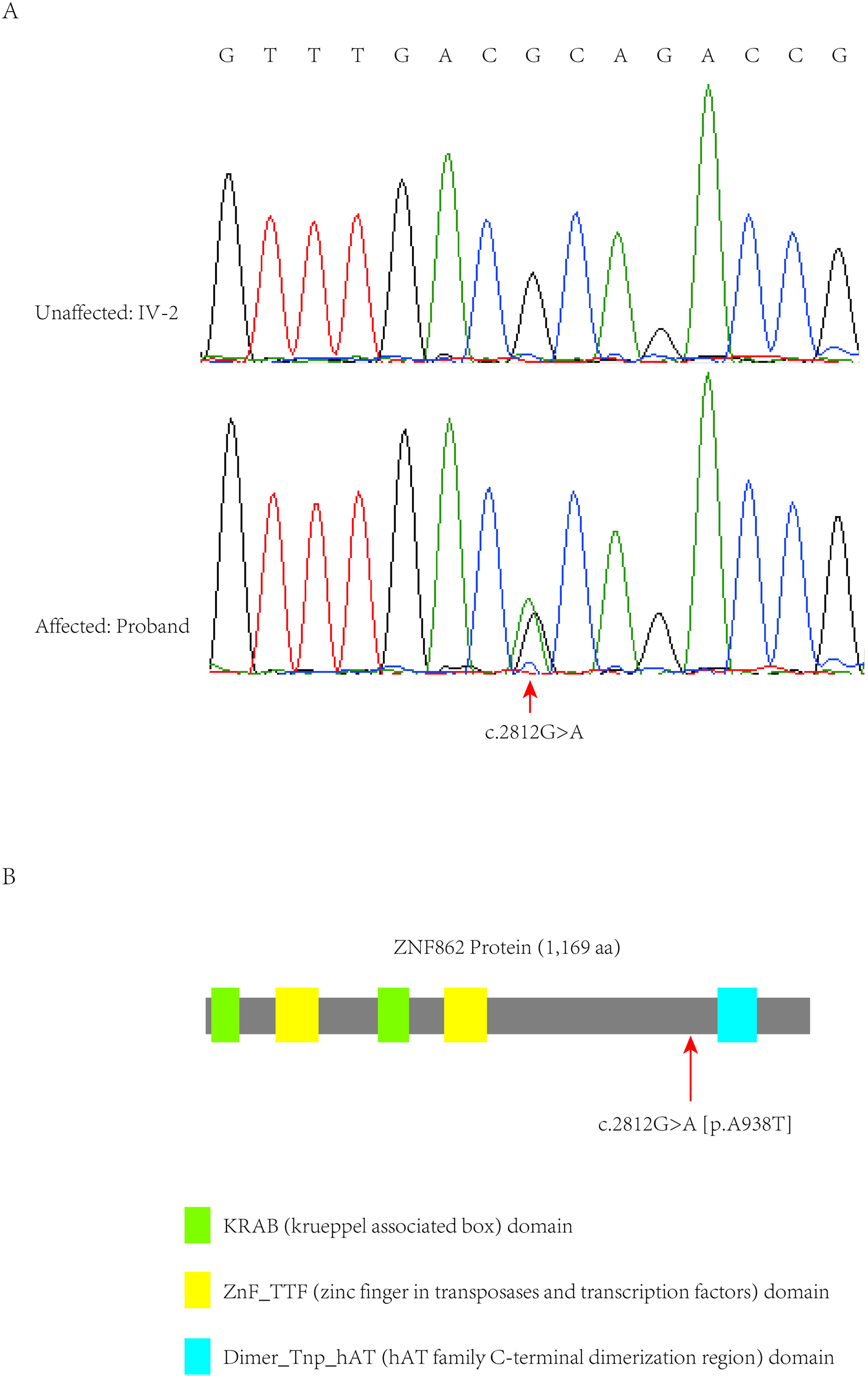
Co-segregation Analysis and the Variant Distribution in the Family. (A) In this family, all affected individuals harbor the heterozygous variant (c.2812G>A) whereas the unaffected individuals are wild-type. (B) Schematic structure with putative domains of ZNF862 protein and the localization of the novel variant (red arrows). Green rectangles indicate KRAB (krueppel associated box) domains; yellow rectangles indicate ZnF_TTF (zinc finger in transposases and transcription factors) domains; blue rectangle indicates Dimer_Tnp_hAT (hAT family C-terminal dimerization region) domain.

*ZNF862* gene harbors 8 exons, encodes a 1,169-amino acid protein, and is located on chromosome 7q36.1. It does not escape our notice that *ZNF862* does not map to any one of the loci that were reported previously as be associated with HGF. ZNF862 is a predicted intracellular protein which function, to the best of our knowledge, is not yet identified, as a zinc finger protein it may be involved in transcriptional regulation. The WES-identified heterozygous variant (p.A938T) in our study located close to and upstream of the Dimer_Tnp_hAT (hAT family C-terminal dimerization region) domain of ZNF862 protein (Figure 2B).

The pathophysiologic mechanisms underlying HGF remain largely elusive. Nevertheless, overproduction of extracellular matrix, particularly the major component collagen type I, may account for the gingival fibroblast overgrowth phenotype; meanwhile increased synthesis of TIMP-1 seem to be associated with collagen I excessive accumulation in HGF fibroblasts.^27,28^ Besides, Martelli-Junior et al. reported that overexpression of TGF-β1 and IL-6 in fibroblasts play pivotal roles in increasing the synthesis of the collagen I along with other specific growth factors.^29^ Systematically, transcriptomic analysis of HGF patients and controls performed by Han et al. demonstrated that regulatory network connection of TGF-β/SMAD signaling pathway and craniofacial development processes contributes to the molecular mechanism of clinical-pathologic manifestations.^30^

Taken together we can speculate that this missense mutation in ZNF862 may increases the collagen synthesis which would result in HGF. ZNF862 and the previously identified protein REST may function as transcriptional factors in a similar way since they both have the zinc-finger DNA-binding domain. Nevertheless, the underlying mechanism of this novel mutation leading to HGF remains enigmatic, with no specific experiment designed for exploring the biological function of the missense variant of ZNF862. Further studies are expected to expand the mutational spectrum of *ZNF862* and the associations with phenotypes, and to investigate the physiological role of ZNF862 in the process of HGF.

In this study, we identify a missense variant (c.2812G>A) in a novel gene *ZNF862*, that causes autosomal-dominant HGF in a four-generation Chinese family, which may shed lights on the precise genetic diagnosis and on the potential therapy development, helping individuals continue their lives better.

## Data Availability

The sequencing data supporting this study have been deposited in the China Genebank Nucleotide Sequence Archive (https://db.cngb.org/cnsa, accession number CNP0000995).

## Data Availability

Data Availability
The sequencing data supporting this study have been deposited in the China Genebank Nucleotide Sequence Archive (https://db.cngb.org/cnsa, accession number CNP0000995).

## Acknowledgments

We thank all the participants in this study, and the support by the funding from National Natural Science Foundation of China (51772144); the Medical Science and Technology Development Foundation, Nanjing Department of Health (YKK18121); Major Project in 13th Five-year of Medical Science and Technology Innovation Platform, Nanjing Department of Health (ZDX16007); Jiangsu Province Natural Science Foundation of China (BK20170143).

D.C., H.H., Y.W., H.C., T.Z., Z.P. and W.L. are employees of BGI Genomics.

## Web Resources

1000 Genomes, http://www.internationalgenome.org/

ClinVar, https://www.ncbi.nlm.nih.gov/clinvar/

Exome Aggregation Consortium (ExAC), http://exac.broadinstitute.org

Genome Aggregation Database, https://gnomad.broadinstitute.org/

GenBank, http://www.ncbi.nlm.nih.gov/genbank/

OMIM, http://www.omim.org/

UniProt, http://www.uniprot.org/

## References

1 Balaji, P. & Balaji, S. M. Gingival fibromatosis with hypertrichosis syndrome: Case series of rare syndrome. Indian J Dent Res 28, 457–460, doi: 10.4103/ijdr.IJDR_367_17 (2017).

2 Snyder, C. H. Syndrome of Gingival Hyperplasia, Hirsutism, and Convulsions; Dilantin Intoxication without Dilantin. J Pediatr 67, 499–502, doi: 10.1016/s0022-3476(65)80413-9 (1965).

3 Witkop, C. J., Jr. Heterogeneity in gingival fibromatosis. Birth Defects Orig Artic Ser 7, 210–221 (1971).

4 Guglielmi, F., Staderini, E., lavarone, F., Di Tonno, L. & Gallenzi, P. Zimmermann-Laband-1 Syndrome: Clinical, Histological, and Proteomic Findings of a 3-Year-Old Patient with Hereditary Gingival Fibromatosis. Biomedicines 7, doi: 10.3390/biomedicines7030048 (2019).

5 Poulopoulos, A., Kittas, D. & Sarigelou, A. Current concepts on gingival fibromatosis-related syndromes. J Investig Clin Dent 2, 156–161, doi:10.1111/j.2041-1626.2011.00054.x (2011).

6 Hakkinen, L. & Csiszar, A. Hereditary gingival fibromatosis: characteristics and novel putative pathogenic mechanisms. J Dent Res 86, 25–34, doi:10.1177/154405910708600104 (2007).

7 Suhanya, J., Aggarwal, C., Mohideen, K., Jayachandran, S. & Ponniah, I. Cherubism combined with epilepsy, mental retardation and gingival fibromatosis (Ramon syndrome): a case report. Head Neck Pathol 4, 126–131, doi:10.1007/s12105-009-0155-9 (2010).

8 Gita, B., Chandrasekaran, S., Manoharan, P. & Dembla, G. Idiopathic gingival fibromatosis associated with progressive hearing loss: A nonfamilial variant of Jones syndrome. Contemp Clin Dent 5, 260–263, doi:10.4103/0976-237X.132387 (2014).

9 Hennekam, R. C. Costello syndrome: an overview. Am J Med Genet C Semin Med Genet 117C, 42–48, doi:10.1002/ajmg.c.10019 (2003).

10 Morey, M. A. & Higgins, R. R. Ectro-amelia syndrome associated with an interstitial deletion of 7q. Am J Med Genet 35, 95–99, doi:10.1002/ajmg.1320350118 (1990).

11 Hamada, K., Ohdo, S., Hayakawa, K., Kikuchi, I. & Koono, M. Puretic syndrome--gingival fibromatosis with hyaline fibromas. Jinrui Idengaku Zasshi 25, 249–255, doi: 10.1007/BF01997703 (1980).

12 Livada, R. & Shiloah, J. Gummy smile: could it be genetic? Hereditary gingival fibromatosis. J Tenn Dent Assoc 92, 23–26; quiz 27–28 (2012).

13 Jorgenson, R. J. & Cocker, M. E. Variation in the Inheritance and Expression of Gingival Fibromatosis. J Periodontol 45, 472–477, doi:10.1902/jop.1974.45.7.472 (1974).

14 Majumder, P., Nair, V., Mukherjee, M., Ghosh, S. & Dey, S. K. The autosomal recessive inheritance of hereditary gingival fibromatosis. Case Rep Dent 2013, 432864, doi: 10.1155/2013/432864 (2013).

15 Xiao, S. et al. A new locus for hereditary gingival fibromatosis (GINGF2) maps to 5q13-q22. Genomics 74, 180–185, doi: 10.1006/geno.2001.6542 (2001).

16 Ye, X. et al. A novel locus for autosomal dominant hereditary gingival fibromatosis, GINGF3, maps to chromosome 2p22.3-p23.3. Clin Genet 68, 239–244, doi:10.1111/j.1399-0004.2005.00488.x (2005).

17 Zhu, Y. et al. A novel locus for maternally inherited human gingival fibromatosis at chromosome 11p15. Hum Genet 121, 113–123, doi:10.1007/s00439-006-0283-1 (2007).

18 Xiao, S. et al. Refinement of the locus for autosomal dominant hereditary gingival fibromatosis (GINGF) to a 3.8-cM region on 2p21. Genomics 68, 247–252, doi: 10.1006/geno.2000.6285 (2000).

19 Hart, T. C. et al. A mutation in the SOS1 gene causes hereditary gingival fibromatosis type 1. Am J Hum Genet 70, 943–954, doi: 10.1086/339689 (2002).

20 Bayram, Y. et al. REST Final-Exon-Truncating Mutations Cause Hereditary Gingival Fibromatosis. Am J Hum Genet 101, 149–156, doi:10.1016/j.ajhg.2017.06.006 (2017).

21 Ahmed, S. & Ali, Z. Rare Case of Idiopathic Gingival Fibromatosis Affecting Primary Dentition. J Ayub Med Coll Abbottabad 27, 933–935 (2015).

22 Gawron, K., Lazarz-Bartyzel, K., Potempa, J. & Chomyszyn-Gajewska, M. Gingival fibromatosis: clinical, molecular and therapeutic issues. Orphanet J Rare Dis 11, 9, doi:10.1186/s13023-016-0395-1 (2016).

23 Odessey, E. A., Cohn, A. B., Casper, F. & Schechter, L. S. Hereditary gingival fibromatosis: aggressive 2-stage surgical resection in lieu of traditional therapy. Ann Plast Surg 57, 557–560, doi: 10.1097/01.sap.0000229059.20539.0e (2006).

24 Chaurasia, A. Hereditary gingival fibromatosis. Natl J Maxillofac Surg 5, 42–46, doi:10.4103/0975-5950.140171 (2014).

25 Zhou, M., Xu, L. & Meng, H. X. Diagnosis and treatment of a hereditary gingival fibromatosis case. Chin J Dent Res 14, 155–158(2011).

26 Zhang, L. et al. Genomic analyses reveal mutational signatures and frequently altered genes in esophageal squamous cell carcinoma. Am J Hum Genet 96, 597–611, doi:10.1016/j.ajhg.2015.02.017 (2015).

27 Gawron, K. et al. TIMP-1 association with collagen type I overproduction in hereditary gingival fibromatosis. Oral Dis 24, 1581–1590, doi:10.1111/odi.12938 (2018).

28 Roman-Malo, L., Bullon, B., de Miguel, M. & Bullon, P. Fibroblasts Collagen Production and Histological Alterations in Hereditary Gingival Fibromatosis. Diseases 7, doi: 10.3390/diseases7020039 (2019).

29 Martelli-Junior, H., Cotrim, P., Graner, E., Sauk, J. J. & Coletta, R. D. Effect of transforming growth factor-beta1, interleukin-6, and interferon-gamma on the expression of type I collagen, heat shock protein 47, matrix metalloproteinase (MMP)-1 and MMP-2 by fibroblasts from normal gingiva and hereditary gingival fibromatosis. J Periodontol 74, 296–306, doi:10.1902/jop.2003.74.3.296 (2003).

30 Han, S. K., Kong, J., Kim, S., Lee, J. H. & Han, D. H. Exomic and transcriptomic alterations of hereditary gingival fibromatosis. Oral Dis 25, 1374–1383, doi: 10.1111/odi.13093 (2019).

